# Socioeconomic and Demographic Barriers Associated with Delays in Pancreatic Cancer Germline Genetic Testing

**DOI:** 10.1101/2023.07.17.23292736

**Authors:** Xianghui Zou, Baho Sidiqi, Sunita Patruni, Leora Rezak, Christopher Hollweg, Noah Kauff, Daniel King

## Abstract

**Background:** Germline genetic testing is recommended for patients with pancreatic ductal adenocarcinoma (PDAC) and pre-diagnostic testing is offered to patients with a significant family history. However, only 41% of patients in our institution obtained genetic testing. We identified associations between patient social profiles and delays in obtaining germline genetic testing from New York’s largest healthcare system.

**Methods:** Patients with PDAC were identified using our EMR between Mar 2016 and Feb 2022 with an IRB-approved protocol. Median income was extrapolated using zip code. Date of diagnosis (DOD) was recorded as the date of biopsy. Delays of testing was calculated as the difference between DOD and the date of germline test. Social work needs and insurance were captured by EMR review of social work notes.

**Results:** 329 patients with PDAC were identified, with 135 (41%) having reports found. Availability of germline testing did not vary by median income. Pearson analysis between income and delays showed a negative correlation (r=-0.258, p=0.0025). Patients who received social security and were unemployed/disabled had significant delays (167d) in testing compared to patients receiving social security (13d) and retired or patients receiving salaries (30d).

African Americans and Hispanics, classified as underserved minority, had significant delays in testing (66d) compared to not underserved patients (22.5d, p=0.021). In addition, African American patients had significant delays in testing (66d) compared to White patients (20d, p=0.0076).

Patients with social work needs had significant delays in testing (104d) compared to patients without SW needs (20.5d, p=0.0002). Of the twelve patients who required SW, six required home care, three required transportation, and two required financial assistance. In addition, patients with Medicare and supplementary insurances had significant decreases in delays (10d) of germline testing compared with patients with Medicare alone (32d, p=0.0077), Medicaid (57d, p=0.020), or commercial insurances (21d, p=0.021).

We identified 12 patients who had germline mutation reports before PDAC diagnosis. For patients with pre-diagnostic testing, 5 had Medicare with supplementary insurances. For the rest of patients with germline testing after the diagnosis of PDAC, 10 had supplementary insurances (p=0.0043).

**Conclusion:** The analysis of our 329-patient cohort showed a striking and concerning negative correlation between patient income and delays in germline testing. Under-represented minority patients had significant delays in germline testing and did not obtain any pre-diagnostic testing. Social work needs and insurance may be barriers as well. Interprofessional collaborations may be required to prompt germline testing.

## Introduction

Germline genetic testing is recommended for all patients with pancreatic ductal adenocarcinoma (PDAC) (1). Approximately 10% of patients with PDAC have inherited pathogenic mutations associated with an increased risk of developing pancreatic cancer (2), and these mutations can serve as predictive biomarkers guiding therapy selection for patients, including initial treatment choice (3), maintenance therapy (4), or other biomarker-determined therapy options (5). Additionally, detection of deleterious mutations should prompt cascade testing in family members, which may save lives through enhanced screening (6). Therefore, expeditious germline mutation profiling is essential to guide optimal management for patients and their families. Despite the importance of germline testing and guidelines recommending its use for patients with PDAC the percentage of patients with PDAC who undergo germline testing remains low, in the range of 21% (7) to 32% (8).

Whereas data explaining low testing rates in pancreatic cancer are lacking, studies of deficiencies in germline testing for prostate cancer uncovered several factors, including lack of follow-up due to transportation issues, higher out-of-pocket costs due to lack of insurance, complicated processes that require frequent visits, patient education and informed consent before the actual germline genetic test (8-11); we hypothesized that patients with PDAC may have similar barriers and sought to quantify the burden of these causes, among others. Additionally, data were lacking describing the punctuality of germline testing in pancreatic cancer, which is crucial, as germline testing results inform optimal first-line treatment. This works presents the prevalence and distribution of delays in germline testing in patients diagnosed with PDAC in our healthcare system (12, 13).

## Materials and Methods

Patients with PDAC were identified using the Northwell Health cancer registry between March 2016 and February 2022 under an IRB-approved protocol. Patient demographics consisting of race, ethnicity, and primary language were queried from data captured during patient registration. Median income was extrapolated using the zip code of the patient’s address and median income public census data (14). Social vulnerability index was collected from the Agency for Toxic Substances and Disease Registry website (15). Insurance data was collected through electronic medical record (EMR) review and grouped into the following categories: Medicare alone, Medicaid, Medicare with supplemental insurance, commercial employee plans, and self-pay. Patient source of income was collected from social work assessment notes and categorized as the following: salary, retired receiving social security or pension, and other due to unemployed or disabled patients.

Germline mutation data were extracted through EMR review of uploaded germline testing reports and by review of reports ordered by Northwell Health oncologists received through request by Invitae®. Diagnosis date was defined as the date of the first histological or cytological confirmation of PDAC. The germline testing date was defined as the sample collection date listed on the germline testing report. Delay in germline testing was calculated as the difference in days between the germline testing date and the diagnosis date. For patients with germline testing before the diagnosis of pancreatic cancer, the delay was recorded as zero days. The social worker assessment note served as the source of social worker needs and patient insurance type. A patient was considered an underserved minority if they self-reported African American race or Hispanic ethnicity. Continuous variables were expressed using sample medians and categorical variables were expressed as percentages. The chi-squared method was used to compare categorical variables. Pearson correlations were used measure the strength of association between continuous variables. All analysis was done using SPSS (IBM SPSS Statistics for Macintosh, Version 24.0, Armonk, NY) using a P value < .05 for statistical significance.

## Results

Overall, 329 patients with PDAC were identified and met the inclusion criteria for data analysis. Germline testing was performed in 135 patients (41.0%). Median income and demographic factors, including sex, race, and ethnicity, were not significantly associated with the performance of germline genetic testing (Table 1).

**Table 1:** Demographic distributions of patient population.

|  |  | n | Performed | Not Performed | p-value |
| --- | --- | --- | --- | --- | --- |
|  |  | 329 | 135 | 194 |  |
| Sex | Male | 140 (43%) | 55 (41%) | 85 (44%) | 0.650 |
|  | Female | 189 (57%) | 80 (59%) | 109 (56%) |  |
| Race | Asian | 29 (9%) | 10 (7%) | 19 (10%) | 0.575 |
|  | African American | 50 (15%) | 19 (14%) | 31 (16%) |  |
|  | White | 185 (56%) | 82 (61%) | 103 (53%) |  |
|  | Others/Unknown | 65 (20%) | 24 (18%) | 41 (21%) |  |
| Ethnicity | Non-Hispanic | 255 (84%) | 119 (88%) | 166 (86%) | 0.105 |
|  | Hispanic | 24 (7%) | 12 (9%) | 12 (6%) |  |
|  | Others/Unknown | 20 (9%) | 4 (3%) | 16 (8%) |  |
| Median Income |  |  | 96318 | 95528 | 0.830 |

Among all tested patients, there was a median delay of 27 days (IQR 10 - 101). There were 12 of 135 patients who had germline testing prior to diagnosis (8.9%). Neither age (r=0.005, p=0.952) nor sex (p=0.497) was associated with delays (Supplemental Fig. 1a-b). Underserved minorities had longer delays, with median time to testing of 66.0 days vs. 22.5 days in nonminority groups (p=0.021) (Fig. 1a). African Americans lad longer delays, of 66.0 days vs. 20.0 days in White patients (p=0.0076). Hispanic patients had numerically longer delays of 84.0 days vs. 27.0 days among the non-Hispanic patients, but was not significant (p=0.481) (Fig. 1b-c).

**Figure 1.**
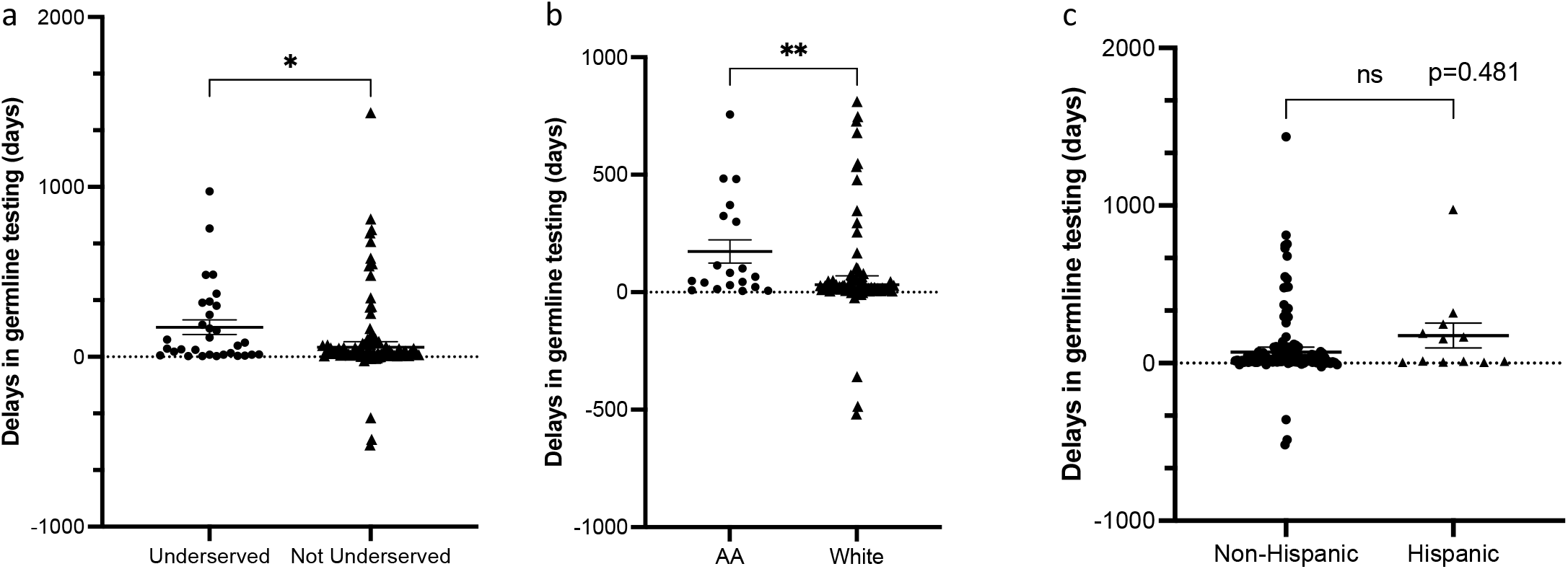
Underserved minorities had significant delays in obtaining germline genetic testing. a) patients were categorized into underserved minorities (African American and Hispanic, n=31) and not underserved (n=104), and their delays in obtaining germline genetic testing were compared using Mann-Whitney test; b) comparison between African American (AA, n=19) and White patients (n=82) regarding delays in obtaining germline genetic testing using Mann-Whitney test; c) comparison between non-Hispanic (n=119) and Hispanic patients (n=12) regarding delays in obtaining germline genetic testing using Mann-Whitney test. * denotes p<0.05, ** denotes p<0.01.

To investigate for causative factors contributing to delays in germline testing, we compared germline testing delay by social work needs and insurance type, as these factors were ascribed to contributing to delays in germline testing rates in other cancer types. In our data set, patients with social work needs had significant delays in germline testing, of 104.0 days vs. 20.5 days in patients without social needs (p=0.0002) (Fig. 2a). Through EMR review, we delineated different categories of social work needs, including need for home care, transportation, and financial assistance (Supplemental Table 1). There was a negative correlation (r=-0.258, p=0.0025) between median income by zip code and delays in testing (Fig. 2b). In addition, there was a positive correlation (r=0.215, p=0.013) between social vulnerability index and delays in testing (Fig. 2c).

**Figure 2.**
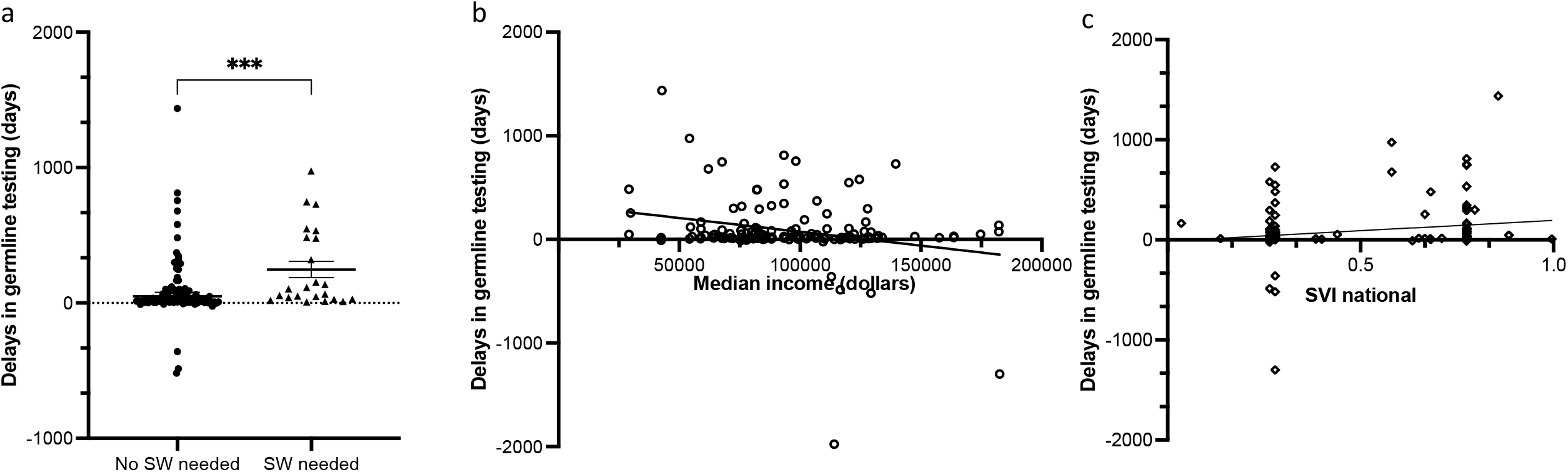
Socioeconomical barriers contribute to delays in germline genetic testing. a) patients were categorized into those who do not need social work (no SW, n=112) and those who need SW (n=23), and their delays in obtaining germline genetic testing were compared using Mann-Whitney test; b) patient median income and delays in obtaining germline genetic testing were plotted in the x- and y-axis, and correlation analysis was performed; c) SVI national index and delays in obtaining germline genetic testing were plotted in the x- and y-axis, and correlation analysis was performed; *** denotes p<0.001.

Patients with primary insurance without any supplemental insurance had significant delays in germline testing. Specifically, patients with Medicare and supplemental insurance had a median delay of 10.0 days compared to 32.0 days for patients with Medicare alone (p=0.0077), 57.0 days for patients with Medicaid alone (p=0.020), and 22.0 days for patients with commercial insurances (p=0.021) (Fig. 3a). In addition to insurance types, testing delays were also associated with source of income (p=0.047) (Fig. 3b), as patients receiving salaries had a median delay of 10.0 days compared to a median delay of 29.0 days for patients receiving social security, pension, or with no source of income. Primary language spoken also showed a numerical difference with non-English speaking patients on median facing 50.0-day delays compared to English speaking patients facing 26.0-day delays (p=0.126) (Fig. 3c).

**Figure 3.**
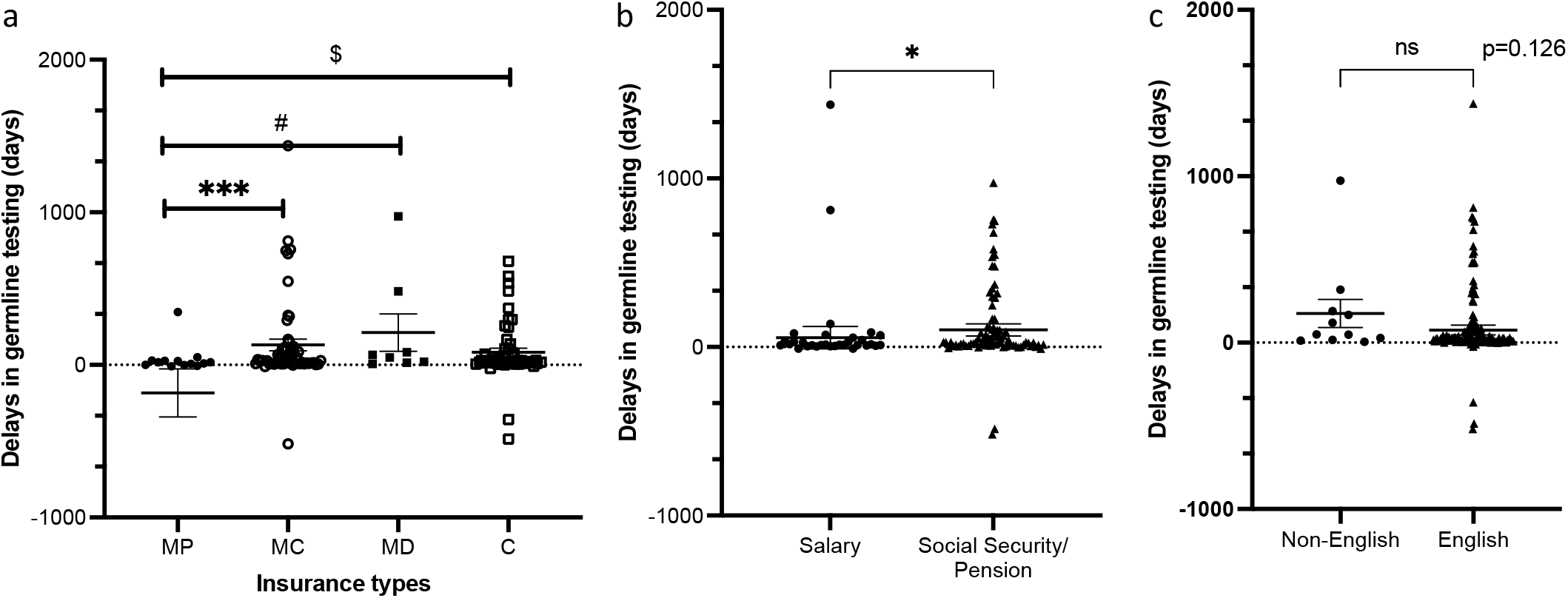
Patient insurances, sources of income, and primary language are associated with delays in germline testing. a) patient insurances were categorized into Medicare with supplemental insurance (MP, n=15), Medicare (MC, n=56), Medicaid (MD, n=8) or Commercial (C, n=51), and Mann-Whitney tests were performed comparing delays between MP and MC, MP and MD, and MP and C; b) patient sources of income were categorized into those who received salaries or those who receive social security or pension, and Mann-Whitney tests were performed to compare delays in obtaining germline genetic testing; c) patient were categorized into those whose primary language is non-English (n=11) and those whose primary language is English (n=123), and Mann-Whitney tests were performed to compare delays in obtaining germline genetic testing; * denotes p<0.05, *** denotes p<0.001 between MP and MC, # denotes p<0.05 between MP and MD, and $ denotes p<0.05 between MP and C.

Among all patients with germline testing, 12 patients had germline mutation testing performed before their biopsy date. Eleven out of these 12 patients had race and ethnicity documented, all of whom were White and non-Hispanic. Among the 5 patients with germline genetic testing predating the diagnosis of pancreatic cancer for more than 100 days, four patients had family or personal history of pancreatic, breast, or colorectal cancers with 3 patients with Medicare plus as their insurance and 2 were Jewish. Overall, for the 12 patients with pre-diagnostic testing, 5 of them had Medicare with supplemental as their insurance, 1 had Medicare/Medicaid and 6 had commercial insurance; whereas for the 123 patients with testing after the diagnosis of pancreatic cancer, 10 of them had Medicare with supplemental as their insurance (p=0.0043) (Supplementary Table 2).

## Discussion

In this study, we have highlighted significant delays in germline genetic testing that are influenced by racial, social, and economic disparities. Among the tested groups, the longest delay observed was a five-fold difference between patients with and without social work needs. This discrepancy likely arises from a combination of factors, including socioeconomic status and various social determinants of health within this population. It has been well-documented that patients facing transportation barriers experience substantial delays in accessing healthcare (16). The lack of sick days in unskilled labor jobs can also prevent timely appointments with a physician (17). Further supporting this evidence, we found that median income is associated with delays in germline testing, reflecting the impact of lower income and limited access to supplementary insurances, particularly in minority populations where Medicare with supplemental coverage is less accessible (18). Additionally, we discovered that underserved minorities face a three-fold increase in delays compared to non-minority patients, with African American patients exhibiting a particularly substantial 3.5-fold increase compared to White patients. This pattern of delay is predominantly observed in underserved minority populations, as consistently noted in multiple studies (8, 19-21). Interventions aimed at improving healthcare literacy in African American communities, such as utilizing a healthcare literacy toolkit to address HIV-related knowledge gaps, have shown promises (22, 23). Similarly, the use of visual learning formats has proven effective in enhancing pancreatic cancer healthcare literacy (24). Our study also revealed a clinically significant linguistic barrier, as non-English-speaking patients often encounter frustrations in medical care, face biases towards interpreter services, and experience discrimination (25). While these factors are associated with delays in germline testing, we believe they can partially explain the causative factors behind such delays. These social and economic barriers can significantly impede timely access to healthcare, including germline testing.

With respect to pre-diagnostic testing, we identified that patients with pre-diagnostic germline genetic testing tended to be White, Jewish, and well insured, suggesting that income and family history are important factors to pre-diagnostic testing. Although the guidelines suggest that pancreatic cancer screening should be considered in higher risk patients with first-degree relatives diagnosed with pancreatic cancer or patients with genetic syndromes associated with an increased risk of pancreas cancer (e.g., Peutz-Jeghers syndrome, CDKN2A mutation, BRCA mutation, ATM mutation, etc.), there are potential financial burdens that may cause either delays in receiving tests or refusals (26).

There are limitations this study. First, it is possible that patients who were transferred to our institution had germline genetic tests performed at outside facilities, but results were not uploaded in our system and therefore we could not accurately calculate the delays in germline testing. In addition, we extrapolated a patient’s income from their home zip-code, which is a surrogate and divergence from the actual income and may have limited external validity. Furthermore, we do not have a large sample size of Hispanic population, which affected the power of our statistical analysis regarding delays in Hispanic patients.

Now that our study has identified associations with delays in germline genetic testing, we plan to study interventions that may reduce barriers associated with delays in germline testing. To address these potential barriers in obtaining germline testing for pancreatic cancer patients and to alleviate health care disparities, our institution has started to expedite germline genetic testing for patients in our multi-disciplinary pancreatic cancer clinic by identifying a navigator who serves as a liaison between patients and medical oncologists/geneticists, facilitates patients consents, and notifies patients for follow-up appointments once germline genetic results are available. As was recently described (27), we have proposed an initiative to collaborate with cancer geneticists in order to mitigate health care literacy, especially in underserved minority group, and to ensure that germline genetic testing can be performed at the first visit after the diagnosis of PDAC. We hope that this institutional-level care plan will be an initiation of mitigating disparities in one aspect of pancreatic-cancer, and will lead to a more global awareness of medical disparities in underserved population, eventually streamlining testing for genetic mutations and optimizing patient-directed treatments.

## Data Availability

All data produced in the present study are available upon reasonable request to the authors

## Acknowledgement

We want to thank Dr. Catherine Alfano and Brooke Tortorella for their assistance in insurance categorization and Dr. Joseph Herman for his expertise in making suggestions when revising this manuscript.

