## Supplementary material for "Socioeconomic and Demographic Barriers Associated with Delays in Pancreatic Cancer Germline Genetic Testing": Supp Fig.1 and Table 2

### Supplementary Figure 1

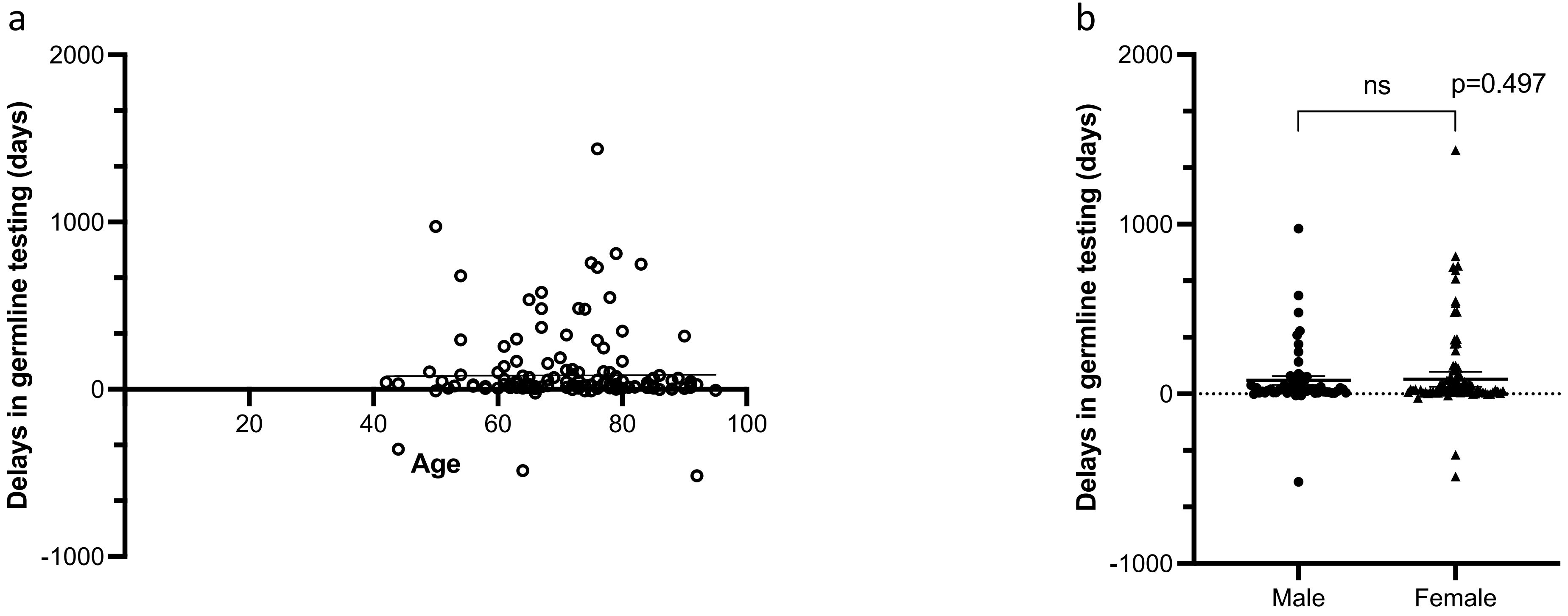


Supplementary Figure 1. **Neither age or gender is associated with delays in germline genetic testing.** a) patient ages and delays in obtaining germline genetic testing were plotted in the x- and y-axis, and correlation analysis was performed; b) male (n=55) and female (n=80) patients were compared regarding delays in obtaining germline genetic testing and Mann-Whitney analysis was performed.

Supplementary Table 2: Comparison of patients with Medicare with supplementary insurances between pre-diagnostic germline testing and post-diagnostic germline testing

| | Medicare with Supplemental insurance | Other insurances | p-value |
|---|---|---|---|
| Pre-diagnostic test | 5 | 7 | 0.0043 |
| Post-diagnostic test | 10 | 113 | |
