## Supplementary material for "Socioeconomic and Demographic Barriers Associated with Delays in Pancreatic Cancer Germline Genetic Testing": Supp Table 1

| Patient numb | Gender | Age range | Zip code | SVI national | Race | Primary Lang |
|---|---|---|---|---|---|---|
| 1 | F | 50-59 | 11423 | 0.7747 | Asian | English |
| 2 | F | 80-89 | 11703 | 0.2645 | White | English |
| 3 | M | 60-69 | 11357 | 0.7747 | White | English |
| 4 | M | 60-69 | 11754 | 0.2645 | White | English |
| 5 | F | 60-69 | 11756 | 0.2782 | Unknown | English |
| 6 | F | 70-79 | 11415 | 0.7747 | White | English |
| 7 | M | 60-69 | 11740 | 0.2645 | Unknown | English |
| 8 | F | 70-79 | 11375 | 0.7747 | White | English |
| 9 | M | 80-89 | 11040 | 0.2782 | White | English |
| 10 | F | 80-89 | 11005 | 0.7747 | White | English |
| 11 | F | 70-79 | 11743 | 0.2645 | White | English |
| 12 | M | 80-89 | 11362 | 0.7747 | White | English |
| 13 | F | 70-79 | 11755 | 0.2645 | White | English |
| 14 | F | 60-69 | 11417 | 0.7747 | Unknown | English |
| 15 | F | 60-69 | 11767 | 0.2645 | White | English |
| 16 | M | 40-49 | 11717 | 0.2645 | Unknown | English |
| 17 | F | 70-79 | 11422 | 0.7747 | African American | English |
| 18 | M | 60-69 | 11565 | 0.2782 | Unknown | English |
| 19 | F | 80-89 | 11791 | 0.2782 | White | Spanish |
| 20 | M | 60-69 | 11355 | 0.7747 | Asian | Cantonese |
| 21 | M | 60-69 | 11365 | 0.7747 | Asian | #N/A |
| 22 | F | 70-79 | 11365 | 0.7747 | Asian | English |
| 23 | F | 70-79 | 11377 | 0.7747 | Asian | English |
| 24 | F | 80-89 | 11362 | 0.7747 | Asian | Korean |
| 25 | F | 80-89 | 11355 | 0.7747 | Unknown | Spanish |
| 26 | F | 60-69 | 10035 | 0.6668 | White | English |
| 27 | F | 70-79 | 11570 | 0.2782 | White | English |
| 28 | F | 70-79 | 11412 | 0.7747 | African American | English |
| 29 | F | 80-89 | 11360 | 0.7747 | White | English |
| 30 | F | 80-89 | 11715 | 0.2645 | White | English |
| 31 | F | 70-79 | 11949 | 0.2645 | White | English |
| 32 | F | 70-79 | 11697 | 0.7747 | White | English |
| 33 | M | 80-89 | 6525 | 0.6512 | White | English |
| 34 | M | 70-79 | 11010 | 0.2782 | Others | English |
| 35 | F | 70-79 | 11758 | 0.2782 | White | English |
| 36 | M | 80-89 | 11040 | 0.2782 | White | English |
| 37 | M | 80-89 | 11378 | 0.7747 | White | English |
| 38 | M | 70-79 | 11373 | 0.7747 | Asian | Mandarin |
| 39 | F | 70-79 | 89109 | 0.8565 | Unknown | English |
| 40 | F | 80-89 | 21044 | 0.1369 | African American | English |
| 41 | F | 80-89 | 11021 | 0.2782 | White | English |
| 42 | F | 50-59 | 10301 | 0.5808 | White | English |

| | | | | | | |
|---|---|---|---|---|---|---|
| 43 | M | 70-79 | 11374 | 0.7747 | Unknown | English |
| 44 | F | 60-69 | 11714 | 0.2782 | White | English |
| 45 | F | 70-79 | 11803 | 0.2782 | White | English |
| 46 | F | 80-89 | 11731 | 0.2645 | Unknown | English |
| 47 | M | 60-69 | 11563 | 0.2782 | White | English |
| 48 | F | 80-89 | 11550 | 0.2782 | Unknown | English |
| 49 | F | 60-69 | 11422 | 0.7747 | African American | English |
| 50 | M | 50-59 | 11731 | 0.2645 | White | English |
| 51 | M | 60-69 | 11040 | 0.2782 | White | English |
| 52 | M | 50-59 | 11414 | 0.7747 | White | English |
| 53 | F | 80-89 | 11377 | 0.7747 | White | English |
| 54 | M | 70-79 | 11426 | 0.7747 | Unknown | Spanish |
| 55 | M | 60-69 | 11545 | 0.2782 | White | English |
| 56 | M | 70-79 | 10562 | 0.6817 | White | English |
| 57 | M | 70-79 | 10520 | 0.6817 | White | English |
| 58 | M | 40-49 | 11434 | 0.7747 | African American | English |
| 59 | M | 70-79 | 11542 | 0.2782 | Unknown | English |
| 60 | M | 50-59 | 11731 | 0.2645 | White | English |
| 61 | F | 80-89 | 11563 | 0.2782 | White | English |
| 62 | M | 70-79 | 11419 | 0.7747 | Asian | English |
| 63 | F | 60-69 | 11797 | 0.2782 | White | English |
| 64 | M | 50-59 | 11545 | 0.2782 | Asian | English |
| 65 | F | 70-79 | 11375 | 0.7747 | African American | English |
| 66 | F | 70-79 | 11429 | 0.7747 | African American | English |
| 67 | M | 80-89 | 11021 | 0.2782 | White | English |
| 68 | M | 70-79 | 11360 | 0.7747 | Asian | English |
| 69 | M | 70-79 | 11758 | 0.2645 | White | English |
| 70 | F | 60-69 | 8801 | 0.0353 | White | English |
| 71 | M | 50-59 | 11368 | 0.7747 | White | English |
| 72 | F | 70-79 | 11714 | 0.2782 | Unknown | English |
| 73 | F | 70-79 | 10465 | 0.9955 | White | English |
| 74 | M | 50-59 | 11414 | 0.7747 | White | English |
| 75 | F | 80-89 | 11377 | 0.7747 | Unknown | Spanish |
| 76 | F | 60-69 | 11720 | 0.2645 | White | English |
| 77 | M | 60-69 | 1590 | 0.4398 | White | English |
| 78 | F | 60-69 | 11105 | 0.7747 | Unknown | English |
| 79 | F | 50-59 | 11239 | 0.8845 | African American | English |
| 80 | M | 60-69 | 11801 | 0.2782 | White | English |
| 81 | F | 70-79 | 11385 | 0.7747 | Unknown | English |
| 82 | F | 60-69 | 11520 | 0.2782 | African American | English |
| 83 | M | 70-79 | 11746 | 0.2645 | African American | English |
| 84 | F | 60-69 | 11716 | 0.2645 | White | English |
| 85 | F | 60-69 | 11362 | 0.7747 | White | English |

| | | | | | | |
|---|---|---|---|---|---|---|
| 86 | F | 70-79 | 11581 | 0.2782 | White | English |
| 87 | M | 80-89 | 11576 | 0.2782 | White | English |
| 88 | F | 70-79 | 11727 | 0.2645 | White | English |
| 89 | F | 60-69 | 11435 | 0.7747 | African American | English |
| 90 | F | 60-69 | 11378 | 0.7747 | White | English |
| 91 | F | 80-89 | 11365 | 0.7747 | White | English |
| 92 | M | 80-89 | 11366 | 0.7747 | White | English |
| 93 | F | 50-59 | 11718 | 0.2645 | White | English |
| 94 | M | 80-89 | 11552 | 0.2782 | White | English |
| 95 | M | 60-69 | 10003 | 0.6668 | White | English |
| 96 | M | 70-79 | 11762 | 0.2645 | White | English |
| 97 | M | 40-49 | 11758 | 0.2645 | White | English |
| 98 | M | 60-69 | 10805 | 0.6801 | White | English |
| 99 | F | 60-69 | 11581 | 0.2782 | White | English |
| 100 | M | 80-89 | 11747 | 0.2645 | White | English |
| 101 | M | 70-79 | 11411 | 0.7747 | African American | English |
| 102 | M | 70-79 | 11428 | 0.7747 | African American | English |
| 103 | F | 80-89 | 11375 | 0.7747 | White | English |
| 104 | F | 70-79 | 11362 | 0.7747 | White | English |
| 105 | M | 50-59 | 10304 | 0.5808 | Others | Spanish |
| 106 | F | 70-79 | 11747 | 0.2645 | Others | English |
| 107 | F | 80-89 | 11419 | 0.7747 | Unknown | Spanish |
| 108 | F | 70-79 | 11411 | 0.7747 | African American | English |
| 109 | M | 70-79 | 18702 | 0.6346 | White | English |
| 110 | F | 70-79 | 11106 | 0.7747 | White | English |
| 111 | M | 60-69 | 7675 | 0.3838 | White | English |
| 112 | F | 60-69 | 7675 | 0.3838 | White | English |
| 113 | F | 50-59 | 8520 | 0.7104 | White | English |
| 114 | M | 80-89 | 11779 | 0.2645 | White | English |
| 115 | F | 60-69 | 11576 | 0.2782 | Unknown | English |
| 116 | M | 60-69 | 11590 | 0.2782 | African American | English |
| 117 | M | 60-69 | 18466 | 0.3994 | Others | English |
| 118 | F | 80-89 | 11742 | 0.2645 | White | English |
| 119 | F | 80-89 | 11362 | 0.7747 | White | English |
| 120 | F | 60-69 | 11010 | 0.2782 | White | English |
| 121 | M | 80-89 | 11004 | 0.7747 | White | English |
| 122 | F | 60-69 | 11229 | 0.8845 | White | Russian |
| 123 | F | 70-79 | 10453 | 0.9955 | African American | English |
| 124 | F | 60-69 | 11542 | 0.2782 | White | English |
| 125 | F | 50-59 | 10596 | 0.6817 | White | English |
| 126 | F | 70-79 | 11793 | 0.2782 | White | English |
| 127 | M | 70-79 | 11776 | 0.2645 | Unknown | Spanish |
| 128 | F | 70-79 | 11364 | 0.7747 | White | English |

| | | | | | | |
|---|---|---|---|---|---|---|
| 129 | F | 80-89 | 11372 | 0.7747 | Unknown | English |
| 130 | M | 80-89 | 11422 | 0.7747 | African American | English |
| 131 | F | 80-89 | 11412 | 0.7747 | African American | English |
| 132 | F | 60-69 | 7036 | 0.7963 | African American | English |
| 133 | F | 60-69 | 11558 | 0.2782 | White | English |
| 134 | F | 40-49 | 11804 | 0.2782 | White | English |
| 135 | F | 60-69 | 11725 | 0.2645 | White | English |

| Insurance | Insurance cat | SW needs | Specific SW | Financial stat | Financial cate | Marital status |
|---|---|---|---|---|---|---|
| Medicaid | MD | No | 0 | Salary | S | Married |
| Medicare/UH | MC | No | 0 | Social Securit | SS | Married |
| GHI NYC EMP | C | No | 0 | Social Securit | SS | Married |
| Magnacare | C | No | 0 | Wage, vetera | S | Single |
| UHC | C | No | 0 | Salary | S | Divorced |
| Medicare | MC | Yes | Home care | Social Securit | SS | Widowed |
| Aetna | C | No | 0 | Social Securit | SS | Married |
| Medicare/GH | MC | No | 0 | NA | NA | NA |
| Medicare/AA | MP | No | 0 | Social Securit | SS | Single |
| Medicare/AA | MP | No | 0 | Social Securit | SS | Widowed |
| Medicare/AA | MP | Yes | Home care | Social Securit | SS | NA |
| Medicare/AA | MP | No | 0 | Social Securit | SS | Married |
| Medicare/BC | MC | No | 0 | Social Securit | SS | Married |
| Healthfirst M | MC | Yes | Home attend | NA | NA | NA |
| BCBS Out of S | C | No | 0 | Social Securit | SS | Married |
| Meritain Hea | C | Yes | Insurance (nc | Unemployed | U | Separated |
| Healthfirst M | MC | No | 0 | Social Securit | SS | Married |
| BCBS MCARE | MC | No | 0 | Salary | S | Divorced |
| Medicare/AA | MP | No | 0 | Social Securit | SS | Widowed |
| Medicare/AA | MP | No | 0 | Social Securit | SS | Divorced |
| UHC | C | Yes | Home care | Salary | S | Married |
| Aetna Mcare | MC | No | 0 | Social Securit | SS | Widowed |
| UHC Medicar | MC | No | 0 | Social Securit | SS | Married |
| Medicare/Me | MC | No | 0 | Social Securit | SS | Widowed |
| Medicare/Me | MC | Yes | Home care | Social Securit | SS | Divorced |
| Magnacare | C | No | 0 | NA | NA | Married |
| BCBS PPO EP( | C | No | 0 | Salary | S | Widowed |
| Aetna Medica | MC | Yes | Ride | Social Securit | SS | Single |
| UHC Medicar | MC | No | 0 | Social Securit | SS | Divorced |
| Medicare | MC | No | 0 | Social Securit | SS | Widowed |
| Medicare/AA | MP | No | 0 | Social Securit | SS | Married |
| Medicare/GH | MC | Yes | Home care, h | Social Securit | SS | Married |
| Medicare/AA | MP | No | 0 | Salary and So | S | Married |
| HIP | C | No | 0 | Social Securit | SS | Married |
| UHC Medicar | MC | Yes | Home care, tı | Social Securit | SS | Widowed |
| Medicare/GH | MC | No | 0 | Social Securit | SS | Married |
| Medicare/AA | MP | No | 0 | Social Securit | SS | Married |
| Healthfirst M | MC | No | 0 | Social Securit | SS | Married |
| Medicare | MC | No | 0 | Salary, retire( | S | Married |
| Medicare/BC | MC | No | 0 | Social Securit | SS | Widowed |
| Medicare | MC | No | 0 | NA | NA | Married |
| GHI NYC EMP | C | No | 0 | Social Securit | SS | Married |

| | | | | | | |
|---|---|---|---|---|---|---|
| Medicare/Tri | MC | No | 0 | Social Securit | SS | Married |
| Medicare/AA | MP | No | 0 | Social Securit | SS | Single |
| UHC Medicar | MC | No | 0 | Social Securit | SS | Divorced |
| Medicare/AA | MP | No | 0 | Social Securit | SS | Married |
| Healthfirst M | MD | Yes | Transportatic | Social Securit | SS | Single |
| Medicare/Me | MC | No | 0 | Social Securit | SS | Widowed |
| Aetna Mcare | C | No | 0 | Social Securit | SS | NA |
| Northwell Dir | C | No | 0 | Salary | S | Married |
| Christian Brot | C | No | 0 | Salary | S | Married |
| UHC Medicar | MC | No | 0 | Salary | S | Married |
| WTC Fund Co | C | No | 0 | Social Securit | SS | Married |
| Healthfirst M | MC | No | 0 | Social Securit | SS | Widowed |
| WTC Fund Co | C | No | 0 | Pension | P | Married |
| NA | NA | Yes | Home care | Social Securit | SS | Divorced |
| UHC Medicar | MC | No | 0 | NA | NA | Married |
| GHI NYC EMP | C | Yes | Financial assi: | Salary | S | Married |
| Humana Med | MC | No | 0 | Pension | P | Married |
| Aetna HMO F | C | No | 0 | Salary | S | Married |
| Medicare | MC | No | 0 | Social Securit | SS | Married |
| HIP Medicare | MC | Yes | Ride | Social Securit | SS | Married |
| Medicare/AA | MP | No | 0 | Salary | S | Married |
| Healthfirst Ex | C | No | 0 | NA | NA | NA |
| Medicare/Me | MC | No | 0 | Social Securit | SS | Married |
| Aetna Mcare | C | No | 0 | Social Securit | SS | Widowed |
| Medicare | MC | No | 0 | Social Securit | SS | Married |
| Wellcare Mca | MC | No | 0 | Social Securit | SS | Married |
| Medicare | MC | No | 0 | Pension and S | P | Married |
| BCBS Out of S | C | No | 0 | Unemployed, | U | Married |
| Aetna HMO | C | No | 0 | Salary | S | Single |
| Medicare/AA | MP | No | 0 | Social Securit | SS | Married |
| UHC Medicar | MC | No | 0 | Social Securit | SS | Unknown |
| BCBS PPO EP( | C | No | 0 | Salary | S | Married |
| Healthfirst M | MC | No | 0 | Social Securit | SS | Married |
| BCBS PPO EP( | C | No | 0 | NA | NA | NA |
| Medicare | MC | No | 0 | Salary | S | Married |
| WTC Fund Co | C | No | 0 | Salary | S | Married |
| NA | NA | No | 0 | NA | NA | NA |
| UHC PPO inde | C | No | 0 | Social Securit | SS | Married |
| UHC Medicar | MC | No | 0 | Social Securit | SS | Single |
| UHC Medicai | MD | No | 0 | Social Securit | SS | Separated |
| Medicare/UH | MC | Yes | Home care | Social Securit | P | Widowed |
| UHC Empire F | C | No | 0 | Salary | S | Married |
| GHI NYC EMP | C | Yes | 911 | Pension and S | P | Single |

| | | | | | | |
|---|---|---|---|---|---|---|
| BCBS Out of S | C | No | 0 | NA | NA | Married |
| Medicare | MC | No | 0 | Social Securit | SS | Married |
| BCBS MCARE | MC | No | 0 | NA | NA | NA |
| Aetna PPO In | C | No | 0 | Salary | S | Married |
| GHI NYC EMP | C | No | 0 | Social Securit | SS | Married |
| Medicare/GH | MC | Yes | Hospital bed | Social Securit | SS | Widowed |
| BCBS MCARE | C | No | 0 | Salary | S | Widowed |
| UHC | C | No | 0 | Social Securit | SS | Married |
| NA | NA | No | 0 | Social Securit | SS | Married |
| Northwell Dir | C | No | 0 | NA | NA | Married |
| Medicare/GH | MC | No | 0 | Social Securit | SS | Married |
| Aetna | C | No | 0 | Salary | S | Married |
| UHC | C | No | 0 | Salary | S | Married |
| Meritain Hea | C | No | 0 | Salary | S | Divorced |
| Medicare/GH | MC | No | 0 | Pension and S | P | Married |
| Medicare | MC | No | 0 | Social Securit | SS | Married |
| Humana Med | MC | No | 0 | Social Securit | SS | Married |
| BCBS MCARE | C | No | 0 | Social Securit | SS | Divorced |
| Aetna Mcare | MC | No | 0 | Salary | S | Divorced |
| Emergency m | MD | Yes | Transportatic | No source of | No income | Separated |
| Medicare/UH | MC | No | 0 | Social Securit | SS | Married |
| HIP Medicare | MC | Yes | Home care | Social Securit | SS | Widowed |
| HIV VIP Medi | MC | No | 0 | Social Securit | SS | Married |
| Medicare/AA | MP | No | 0 | Salary | S | Married |
| HIP Medicare | MC | No | 0 | Social Securit | SS | Widowed |
| Oxford Libert | C | No | 0 | Salary | S | Single |
| Insured throu | C | No | 0 | Salary | S | Married |
| Cigna | C | No | 0 | Social Securit | SS | Divorced |
| Aetna Mcare | C | No | 0 | Social Securit | SS | Widowed |
| BCBS Out of S | C | Yes | Home rehab | Salary | S | Married |
| UHC HMO PC | C | No | 0 | Social Securit | P | Married |
| BCBS HMO P( | C | No | 0 | NA | NA | NA |
| UHC Medicar | MC | No | 0 | NA | NA | NA |
| UHC Medicar | MC | No | 0 | Pension | P | Widowed |
| Local 1199 | C | No | 0 | NA | NA | NA |
| Medicare/AA | MP | No | 0 | Social Securit | SS | Married |
| Fideles Medic | MD | Yes | Home care | Unemployed | U | Widowed |
| GHI NYC EMP | C | Yes | Ride and hom | NA | NA | Single |
| Healthfirst M | MD | No | 0 | Salary | S | Married |
| NA | NA | No | 0 | Salary | S | Married |
| Medicare | MC | Yes | Home care ar | Unemployed | U | Married |
| Self pay unin: | Self pay | No | 0 | Self pay | NA | NA |
| Medicare/UH | MC | No | 0 | NA | NA | NA |

| | | | | | | | |
|---|---|---|---|---|---|---|---|
| UHC Medicar | MC | No | | 0 | NA | NA | NA |
| Medicaid/HIF | MD | No | | 0 | Social Securit | SS | Married |
| Medicaid/HIF | MD | Yes | Hospice | | Social Securit | SS | Widowed |
| Aetna HMO | C | No | | 0 | Social Securit | SS | Single |
| Oxford Freed | C | No | | 0 | Salary | S | Married |
| Magnacare | C | No | | 0 | NA | NA | NA |
| BCBS Out of S | C | No | | 0 | NA | NA | Married |

| g of Diagnosis | Ethnicity | Median Income | Result, pathogenic |
|---|---|---|---|
| 14d | Non-Hispanic | 71194 | no mutations |
| 13d | Non-Hispanic | 93938 | CTFR c.1210-34TG |
| 29d | Non-Hispanic | 85818 | no mutations |
| 35d | Non-Hispanic | 101710 | no mutations |
| 3 | Non-Hispanic | 125486 | PALB2 c.2167_216 |
| 55d | Non-Hispanic | 70900 | no mutations |
| 580d | Unknown | 124527 | no mutations |
| 6d | Non-Hispanic | 84713 | no mutations |
| 10 | Non-Hispanic | 129344 | no mutations |
| 28 | Non-Hispanic | 55313 | no mutations |
| 27d | Non-Hispanic | 128187 | no mutations |
| 347d | Non-Hispanic | 93333 | CFTR c.1210-34TG |
| -1 | Non-Hispanic | 116788 | no mutations |
| 154d | Hispanic | 76868 | no mutations |
| -486d | Non-Hispanic | 116599 | no mutations |
| 104d | Non-Hispanic | 82237 | no mutations |
| 325d | Non-Hispanic | 88130 | no mutations |
| 8d | Non-Hispanic | 128301 | no mutations |
| 50d | Non-Hispanic | 174675 | no mutations |
| 17 | Non-Hispanic | 42507 | no mutations |
| 10 | Non-Hispanic | 67642 | no mutations |
| 90d | Non-Hispanic | 67642 | no mutations |
| 101d | Non-Hispanic | 58948 | no mutations |
| 28d | Non-Hispanic | 93333 | no mutations |
| 12d | Hispanic | 42507 | no mutations |
| 256d | Non-Hispanic | 29799 | no mutations |
| 12d | Non-Hispanic | 117107 | no mutations |
| 114d | Non-Hispanic | 81608 | no mutations |
| 67d | Non-Hispanic | 83063 | no mutations |
| 2 | Non-Hispanic | 122639 | no mutations |
| 25d | Non-Hispanic | 96505 | no mutations |
| 28 | Non-Hispanic | 105750 | no mutations |
| 17d | Non-Hispanic | 157610 | no mutations |
| 247d | Hispanic | 111135 | no mutations |
| 548 | Non-Hispanic | 120149 | no mutations |
| -518d | Non-Hispanic | 129344 | no mutations |
| -6d | Non-Hispanic | 77710 | no mutations |
| 119 | Non-Hispanic | 54584 | no mutations |
| 1437d | Non-Hispanic | 42837 | no mutations |
| 13 | Non-Hispanic | 106533 | no mutations |
| 11d | Non-Hispanic | 102596 | no mutations |
| 679d | Non-Hispanic | 62052 | no mutations |

| | | | | |
|---|---|---|---|---|
| 26d | Non-Hispanic | 66795 | no mutations | |
| -1976d | Non-Hispanic | 114095 | no mutations | |
| 29d | Non-Hispanic | 147469 | no mutations | |
| 6d | Non-Hispanic | 120619 | no mutations | |
| 20 | Non-Hispanic | 99947 | no mutations | |
| 14d | Hispanic | 64155 | no mutations | |
| 5d | Non-Hispanic | 88130 | no mutations | |
| 27d | Non-Hispanic | 120619 | no mutations | |
| 70 | Non-Hispanic | 129344 | no mutations | |
| 86 | Non-Hispanic | 75471 | no mutations | |
| 20 | Non-Hispanic | 58948 | no mutations | |
| 5d | Hispanic | 98987 | no mutations | |
| 30d | Non-Hispanic | 163606 | no mutations | |
| 479 | Non-Hispanic | 81952 | no mutations | |
| 5d | Non-Hispanic | 110094 | no mutations | |
| 41d | Non-Hispanic | 64087 | no mutations | |
| 12d | Hispanic | 80607 | no mutations | |
| 16d | Non-Hispanic | 120619 | no mutations | |
| 16 | Non-Hispanic | 99947 | no mutations | |
| 51d | Non-Hispanic | 75835 | no mutations | |
| -1299d | Non-Hispanic | 182569 | no mutations | |
| 19 | Non-Hispanic | 163606 | no mutations | |
| 44d | Non-Hispanic | 84713 | no mutations | |
| 22d | Non-Hispanic | 88300 | no mutations | |
| 7d | Non-Hispanic | 102596 | no mutations | |
| 292d | Non-Hispanic | 83063 | no mutations | |
| 105d | Non-Hispanic | 120149 | no mutations | |
| 167d | Non-Hispanic | 127130 | BRCA2 c.7617+2T> | |
| 5d | Hispanic | 54211 | no mutations | |
| 0ms | Non-Hispanic | 114095 | no mutations | |
| 10 | Non-Hispanic | 75617 | no mutations | |
| -9d | Non-Hispanic | 75471 | no mutations | |
| 167d | Hispanic | 58948 | no mutations | |
| -23d | Non-Hispanic | 109550 | no mutations | |
| 55 | Non-Hispanic | 104710 | no mutations | |
| 72 | Non-Hispanic | 79883 | no mutations | |
| 48d | Non-Hispanic | 29172 | no mutations | |
| 21d | Non-Hispanic | 110302 | ATM pR250* | Cl |
| -10d | Non-Hispanic | 75340 | no mutations | |
| 482d | Non-Hispanic | 82200 | no mutations | |
| 8 | Non-Hispanic | 116560 | no mutations | |
| 13d | Non-Hispanic | 93317 | no mutations | |
| 535 | Non-Hispanic | 93333 | no mutations | |

| | | | |
|---|---|---|---|
| 18 | Non-Hispanic | 117200 | no mutations |
| 74d | Non-Hispanic | 182199 | no mutations |
| 10d | Non-Hispanic | 82655 | no mutations |
| 30d | Non-Hispanic | 66137 | no mutations |
| 61d | Non-Hispanic | 77710 | no mutations |
| 748d | Non-Hispanic | 67642 | no mutations |
| 5d | Non-Hispanic | 84301 | no mutations |
| 296d | Non-Hispanic | 127931 | no mutations |
| 52d | Non-Hispanic | 122367 | no mutations |
| 15 | Non-Hispanic | 118161 | no mutations |
| 35d | Non-Hispanic | 132426 | no mutations |
| 33d | Non-Hispanic | 120149 | no mutations |
| 11d | Non-Hispanic | 72138 | no mutations |
| 11d | Non-Hispanic | 117200 | no mutations |
| 7 | Non-Hispanic | 126607 | #N/A |
| 101d | Non-Hispanic | 98207 | no mutations |
| 6 | Non-Hispanic | 78368 | no mutations |
| 1d | Non-Hispanic | 84713 | no mutations |
| 812 | Non-Hispanic | 93333 | APC I1307K |
| 974d | Hispanic | 54199 | no mutations |
| 76d | Unknown | 126607 | no mutations |
| 319d | Hispanic | 75835 | no mutations |
| 756d | Non-Hispanic | 98207 | no mutations |
| -9d | Non-Hispanic | 42472 | no mutations |
| 9 | Non-Hispanic | 68530 | ATM N2133Hfs*3 |
| 13d | Non-Hispanic | 131940 | no mutations |
| 10d | Non-Hispanic | 131940 | no mutations |
| 18 | Non-Hispanic | 93419 | no mutations |
| 28d | Non-Hispanic | 98450 | no mutations |
| 137d | Non-Hispanic | 182199 | no mutations |
| 371d | Non-Hispanic | 106959 | no mutations |
| 7d | Hispanic | 58771 | no mutations |
| 45d | Non-Hispanic | 107056 | no mutations |
| 24 | Non-Hispanic | 93333 | no mutations |
| 101d | Non-Hispanic | 111135 | no mutations |
| -1d | Non-Hispanic | 95436 | no mutations |
| 48 | Non-Hispanic | 64631 | no mutations |
| 484 | Non-Hispanic | 29136 | no mutations |
| 6d | Non-Hispanic | 80607 | PALB2 R753* |
| 7d | Non-Hispanic | 85417 | CDKN2A G101W M |
| 729 | Non-Hispanic | 139543 | no mutations |
| 189d | Hispanic | 101793 | no mutations |
| 13d | Non-Hispanic | 82028 | no mutations |

| | | | |
|---|---|---|---|
| 40 | Unknown | 60659 | no mutations |
| 83d | Non-Hispanic | 88130 | no mutations |
| 66 | Non-Hispanic | 81608 | no mutations |
| 300d | Non-Hispanic | 72432 | no mutations |
| 80d | Non-Hispanic | 96368 | APC I1307K |
| -359d | Non-Hispanic | 112946 | no mutations |
| 21d | Unknown | 133893 | no mutations |

Result, VUS
ATM I2316F CTNNA1 A112T
no mutations
no mutations
no mutations
#N/A
no mutations
BARD1 S37G
no mutations
#N/A
#N/A
no mutations
KIT H101Y
#N/A
no mutations
no mutations
ATM D817N
no mutations
no mutations
no mutations
no mutations
no mutations
no mutations
no mutations
no mutations
MET T222M POLE S692L
BRCA1 E1694K
BMPR1 E472D RECQL4 L566P
ALK P1032S ATM C2488Y
no mutations
#N/A
CASR C851S
#N/A
POLE E1424K
no mutations
#N/A
ATM F1977S
no mutations
#N/A
no mutations
#N/A
no mutations
RAD50 pK543_D544insARTNMEMLTK

no mutations
no mutations
no mutations
no mutations
#N/A
POLE A629S
APC non-coding NTHL1 A246T
no mutations
#N/A
#N/A
#N/A
PMS2 R799W
BRCA2 E2599K
#N/A
no mutations
no mutations
no mutations
no mutations
#N/A
TSC1 E860K
no mutations
#N/A
no mutations
NF1 N1430S
no mutations
MSH3 F709L RAD50 N737E
no mutations
no mutations
no mutations
no mutations
#N/A
MSH6 E1272D RAD L172F
no mutations
CHEK2 P182T
#N/A
no mutations
no mutations
no mutations
BRCA I1808T PALB2 A968G
ATM G1746R MSH6 L290P
#N/A
AXIN2 V539M BMPR1A R494N
#N/A

#N/A
no mutations
MSH6 P1087H
BRIP1 Q608H
RAD K264E
DICER1 E512G
no mutations
no mutations
no mutations
#N/A
no mutations
no mutations
BRCA2 S1743N
no mutations
#N/A
no mutations
#N/A
no mutations
AXIN2 pM5R
no mutations
ATM D851E HRAS R169W MEN1 P188L TP53 R283C
BLM intronic mutation
AIP V195A
no mutations
#N/A
no mutations
no mutations
#N/A
BRIP1 R356N
no mutations
DIS3L2 P612L PTCH1 D635N
POLD1 K468R SDHA R589G
no mutations
ATM R1575H
TSC2 R93W
no mutations
#N/A
#N/A
CHEK2 N446D
no mutations
#N/A
no mutations
no mutations

| | |
|---|---|
| MET N1099S | |
| no mutations | |
| | #N/A |
| MSH6 D217G | |
| no mutations | |
| no mutations | |
| no mutations | |
